# A Novel Truncating Variant in MYBPC3 Causes Hypertrophic Cardiomyopathy

**DOI:** 10.1101/2024.02.18.24302943

**Authors:** Yuanyuan Zhang, Wenyan Gong, Yusheng Cong, Xingwei Zhang, Zhelan Zheng

## Abstract

**Background:** Familial hypertrophic cardiomyopathy (HCM) is the most common genetic cardiovascular disease. Related mutations contributing to hypercontractility and poor relaxation in HCM have been incompletely understood. The purpose of this study was to explore and verify a novel variant in cardiac myosin-binding protein C3 (MYBPC3) in a HCM family.

**Methods:** Clinical information was collected and cardiac evaluation was performed in the pedigree. Second-generation sequencing technology was used to investigate the proband and his family. Computational prediction of mutation effects at genomic level and 3D visualization of the mutated protein were achieved by in silico analysis.

**Results:** Typical interventricular septal thickening was detected in all the four HCM patients. A c.1042_1043insCGGCA mutation of MYBPC3 was verified in the proband and family members. Mild phenotype associated with delayed onset and relative favorable prognosis were observed in the pedigree. In silico analysis of the mutation revealed that c.1042_1043insCGGCA led to an early termination of MYBPC protein synthesis at C2 domain, losing the domains that are essential for myosin-and titin-binding.

**Conclusion:** The novel c.1042_1043insCGGCA mutation of MYBPC3 was a genetic basis for HCM.

Our gene sequence based computational analysis predicted the pathogenicity of the mutation by correlating MYBPC3 genotypes with clinical phenotypes.

## 1. Introduction

Hypertrophic cardiomyopathy is an autosomal genetic disease occurred in absence of common triggers such as hypertension and aortic stenosis[1]. The prevalence of HCM is 1:500 in the general population[2]. Striking cardiomegaly and stunning asymmetry with disproportionate involvement of interventricular septum was observed in many HCM cases[3]. Postmortem examination in hearts of HCM victims show myocyte disarray accompanied with significantly increased myocardial fibrosis[4].

Many patients with HCM experience adverse clinical outcomes including heart failure (HF), arrhythmias, and sudden cardiac death. The mortality of patients with HCM was shown to be about 3-fold higher than that of the general population at similar ages[5]. It is the most common cause of sudden cardiac death among young athletes without precedent symptoms.

Many efforts have been made to gain an accurate and comprehensive understanding of the molecular basis and clinical course of the disease. Pathogenic variants in genes encoding sarcomere proteins are involved in hypertrophic cardiomyopathy[6]. More than 64 genes encoding thick and thin filaments have been claimed to be causative in HCM with varying levels of supportive evidence[7].

Here we report a novel mutation in the myosin binding protein from a sporadic pedigree. We use DNA sequencing technology and bioinformatic tools to illustrate the genetic background and possible influence of the mutated protein. We aim to gain evidence upon the relationship between genotype and clinical manifestation, underscoring the value of genomics in elucidating the underlying mechanism of HCM.

## 2. Materials and Methods

### 2.1 Ethical approval

The study was conducted in accordance with the Declaration of Helsinki and was approved by the Ethics Committee of the First Affiliated Hospital of Zhejiang University. Starting on 15 July 2022, the recruitment period ended on 15 March 2023. Written consent was obtained from every participant in the study. A flowchart of the study was shown in Figure 1.

**Figure 1.**
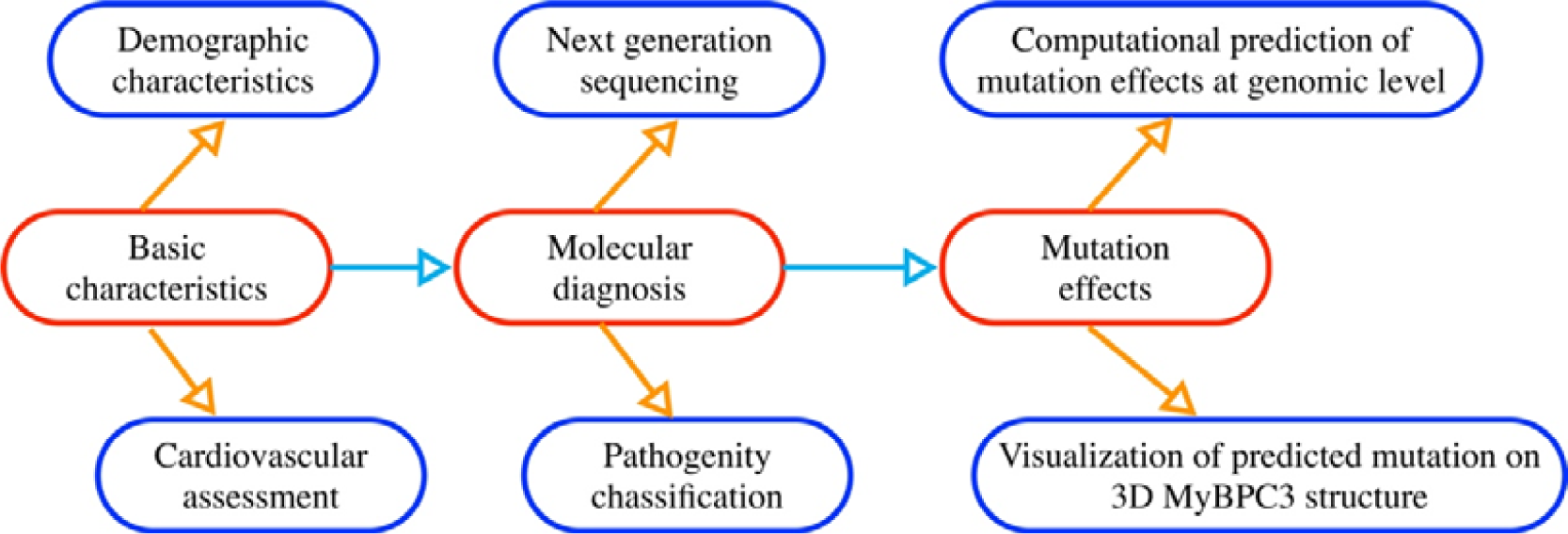
Flow diagram of the study.

### 2.2 Index family

Demographic data was collected for every patient at the day of admission. All patients underwent a complete cardiovascular evaluation, including cardiac physical examination, blood test of brain natriuretic peptide, 12-lead electrocardiogram (ECG) and echocardiography. The diagnostic criteria for hypertrophic cardiomyopathy were according to the recommendations by the American College of Cardiology/American Heart Association Joint Committee [8].

### 2.3 Sample collection and DNA isolation

Genomic DNA from blood samples collected in ethylenediaminetetraacetic acid (EDTA) tubes was extracted using the DNeasy Blood & Tissue Kit (Cat No.69504, Qiagen, Germany) according to the manufacturer’s protocol. The concentration of genomic DNA was determined with an UV-Vis/florescence spectrophotometer (ES-2, MALCOM, Japan).

### 2.4 Genome sequencing

Genomic DNA extracted from peripheral blood was fragmented to an average size of ∼350bp and subjected to DNA library creation using established Illumina paired-end protocols. The Illumina Novaseq 6000 platform (Illumina Inc., San Diego, CA, USA) was utilized for genomic DNA sequencing in Novogene Bioinformatics Technology Co., Ltd (Beijing, China) to generate 150-bp paired-end reads with a minimum coverage of 10× for ∼99% of the genome (mean coverage of 30×). After sequencing, basecall files conversion and demultiplexing were performed with bcl2fastq software (Illumina). The resulting fastq data were submitted to in-house quality control software for removing low quality reads, and then were aligned to the reference human genome (hs37d5) using the Burrows-Wheeler Aligner (BWA) [9], and duplicate reads were marked using Sambamba tools [10]. Genomic variations were annotated using ANNOVAR software [11].

### 2.5 Computational prediction of mutation effects at genomic and protein level

To study the mutation effects on gene regulation at genomic level, DNAMAN V6.0 software was used to carry out multiple sequence alignments. To illustrate the stability and conformational effects on protein level, 3D model was created using online automated protein structure server SWISS-MODEL (https://swissmodel.expasy.org).

## 3 Results

### 3.1 Basic characteristics of the HCM family

Partial pedigree of the family was shown in Figure 2 and main clinical features was shown in Table 1. Briefly, the proband was an old man who was diagnosed with HCM at his 50s. He suffered from repeated episodes of heart failure and was given diuretic drugs. Individual II-2 was diagnosed with HCM at his 50s due to chest discomfort and palpitation. He was given beta-blocker to relieve symptoms. Of note, individual II-3 and II-4 remained asymptomatic thus far, without any episodes of palpitations, arrhythmia, or syncope.

**Figure 2.**
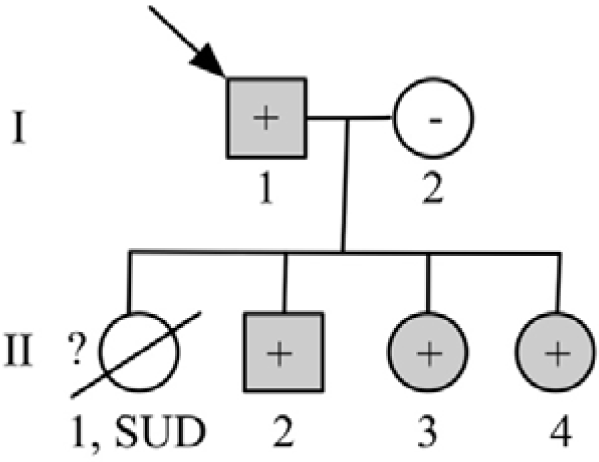
Partial pedigree chart of the HCM family. Each individual is identified with a number and generations are indicated in the left side. Patients with hypertrophic cardiomyopathy are shown in grey, unaffected individuals are shown in white, and slashes indicate a deceased relative. Index case is indicated with an arrow (I-1). Question mark indicates no genetic analysis available. Minus and plus symbols, respectively, represent genotype negative or positive subjects. SUD means sudden unexplained death.

**Table 1.** Baseline characteristics of the HCM patients.

| Demographic characteristics | I-1 | II-2 | II-3 | II-4 |
| --- | --- | --- | --- | --- |
| Sex | Male | Male | Female | Female |
| Age range at diagnosis, y | 50-55 | 50-55 | 50-55 | 45-50 |
| Medical history |  |  |  |  |
| Current smoker | No | No | No | No |
| Diabetes | No | No | No | No |
| Hypertension | Yes | Yes | No | No |
| Dyspnea or shortness of breath | Yes | No | No | No |
| Angina pectoris (chest pain) | No | No | No | No |
| Syncope | No | No | No | No |
| Cardiac arrest | No | No | No | No |
| Surgical myectomy | No | No | No | No |
| Heart failure episodes | Yes | No | No | No |
| NYHA class | II | I | I | I |
| Medications |  |  |  |  |
| Beta blockers | Yes | Yes | Yes | No |
| Calcium antagonists | No | No | No | No |
| Diuretics | Yes | No | No | No |
| Warfarin | No | No | No | No |
| Biomarkers |  |  |  |  |
| BNP (pg/ml) | 434.97 | <35 | <35 | <35 |

Abbreviation: BNP = brain natriuretic peptide; HCM = hypertrophic cardiomyopathy; NYHA = New York Heart Association.

### 3.2 Cardiovascular evaluation of the family

Typical echocardiographic images, along with ECG results were shown in Figure 3 and Figure 4, respectively. Notably, septal thickness increased significantly in all the four patients with inter-ventricular septum dimension (IVSd)/left ventricular posterior wall dimension (LVPWd) ratio > 1.3, indicating the diagnosis of HCM[8,12]. No left ventricular out flow obstruction was detected in the affected members. For echocardiographic details see Table 2. ECG shows significant alterations of the hypertrophic heart. The index case shows increased QRS voltages as SV1 + RV5 and RV6 both are larger than 35 mm. Meanwhile, arrhythmia including atrial premature beat and intraventricular block occurred in the proband. T-wave inversion in at least two adjacent precordial leads was detected in individual II-2, II-3 and II-4.

**Figure 3.**
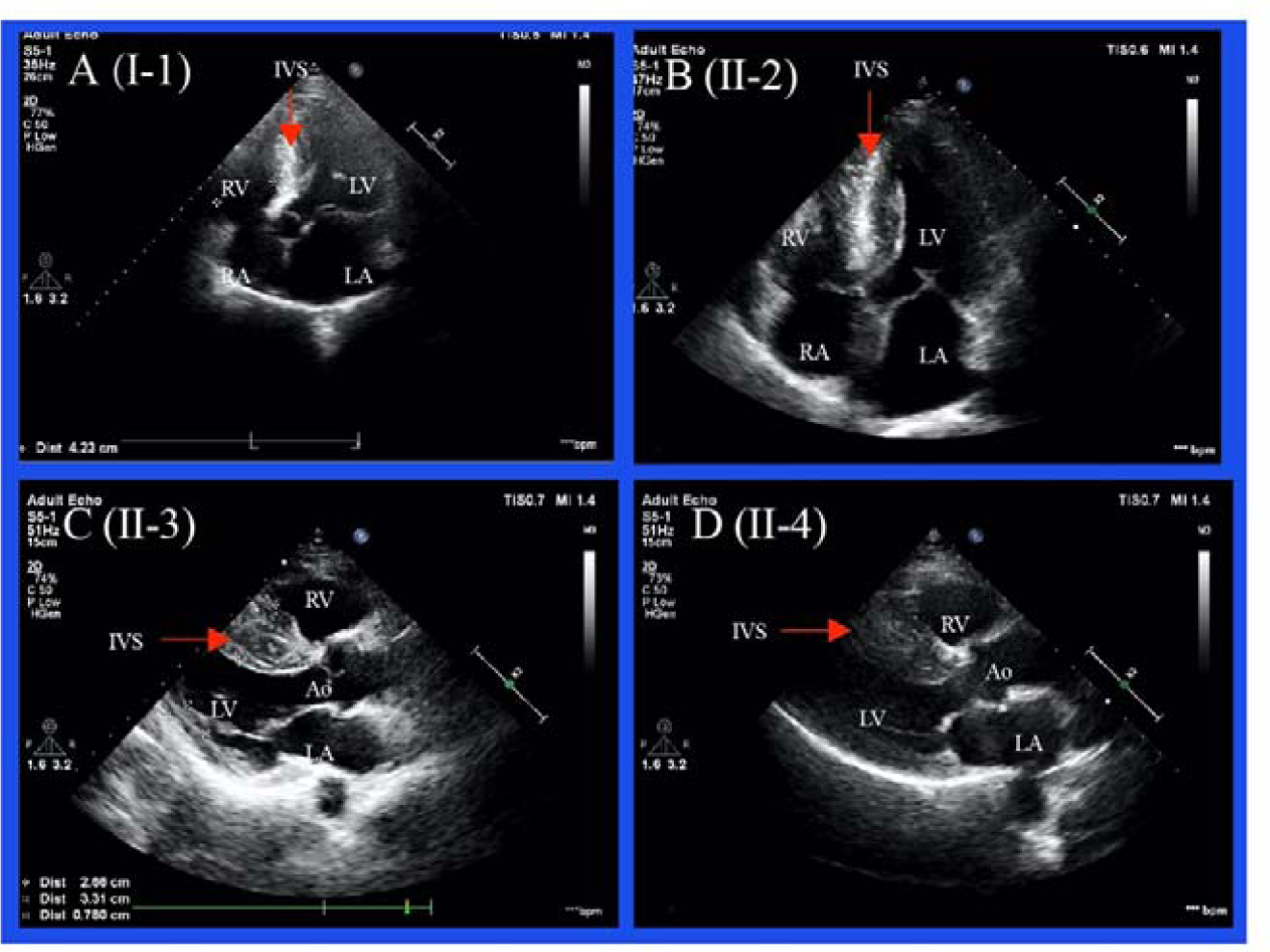
Typical echocardiography of family members. Four chamber view of I-1 and II-2. Parasternal long axis view of II-3 and II-4. LA = left atrium; LV = left ventricle; RA = right atrium; RV = right ventricle; Ao = aorta.

**Figure 4.**
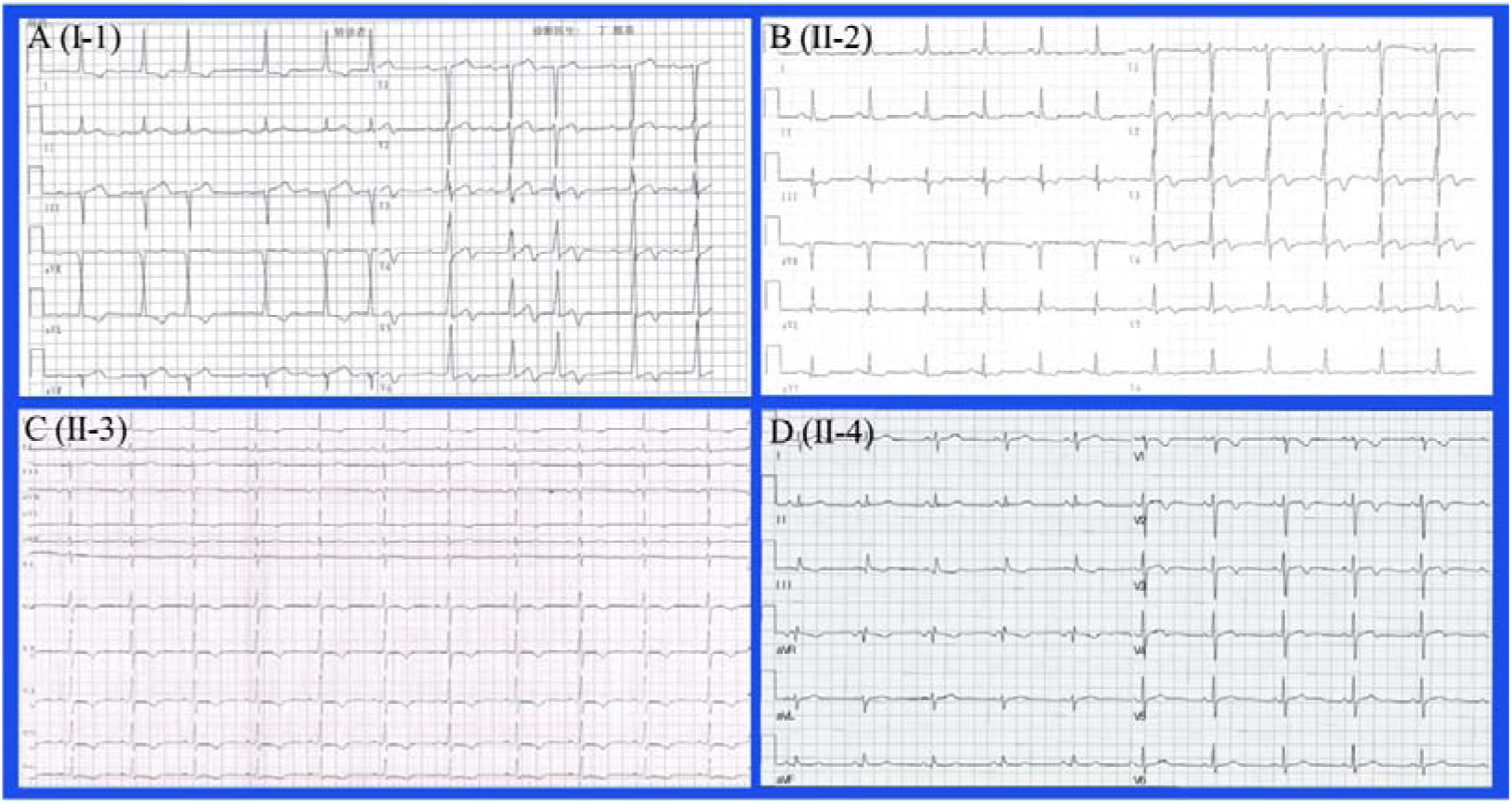
Typical ECG results show significant alterations of the hypertrophic heart.

**Table 2.**
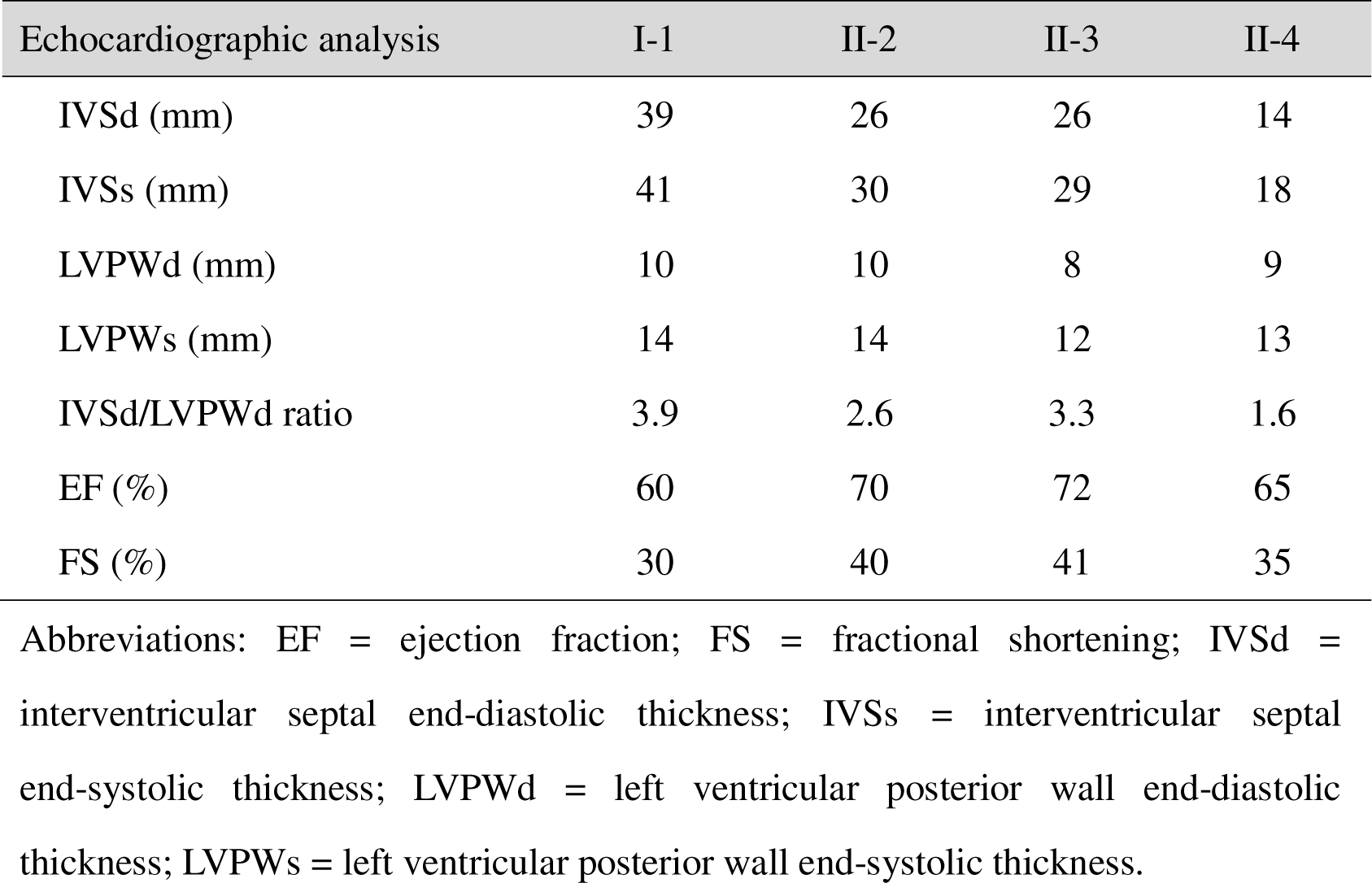
Variable of transthoracic echocardiography of the pedegree.

| Echocardiographic analysis | I-1 | II-2 | II-3 | II-4 |
| --- | --- | --- | --- | --- |
| IVSd (mm) | 39 | 26 | 26 | 14 |
| IVSs (mm) | 41 | 30 | 29 | 18 |
| LVPWd (mm) | 10 | 10 | 8 | 9 |
| LVPWs (mm) | 14 | 14 | 12 | 13 |
| IVSd/LVPWd ratio | 3.9 | 2.6 | 3.3 | 1.6 |
| EF (%) | 60 | 70 | 72 | 65 |
| FS (%) | 30 | 40 | 41 | 35 |

Abbreviations: EF = ejection fraction; FS = fractional shortening; IVSd = interventricular septal end-diastolic thickness; IVSs = interventricular septal end-systolic thickness; LVPWd = left ventricular posterior wall end-diastolic thickness; LVPWs = left ventricular posterior wall end-systolic thickness.

### 3.3 Molecular diagnosis and pathogenicity classification

Mutation screening revealed a candidate mutation in MYBPC3 from all the four patients. A heterozygous mutation c.1042_1043insCGGCA located in exon 12 occurred in the proband and his offspring. Base sequence of exon 12 before and after mutation was shown in the supplementary material. The c.1042_1043insCGGCA mutation causes a shift in the reading frame beginning with Proline, changing it to an Arginine, and creates a premature stop codon at the new reading frame. The amino acid sequence prior and post mutation was shown in the supplementary material. This mutation is expected to result in an abnormal, truncated protein or loss of protein from this allele due to nonsense-mediated mRNA decay.

According to the American College of Medical Genetics (ACMG) framework, the c.1042_1043insCGGCA in the MYBPC3 gene is interpreted as a pathogenic mutation based on the following criteria: PS4: The prevalence of the variant in affected individuals is significantly increased compared to the prevalence in population controls; PM1: The variant located in a mutational hot spot without benign variation; PM2: The variant is at extremely low frequency in the wider population; PM4: Protein length changes due to insertions in a non-repeat region[13].

### 3.4 Prediction of mutant effects on gene regulation and visualization of mutated MYBPC3 protein

In silico analysis of the mutation revealed that c.1042_1043insCGGCA led to an early termination of MYBPC protein synthesis at C2 domain in homo sapiens. The truncated protein ended with aa.350, losing the domains that are essential for myosin-and titin-binding (Figure 5).

**Figure 5.**
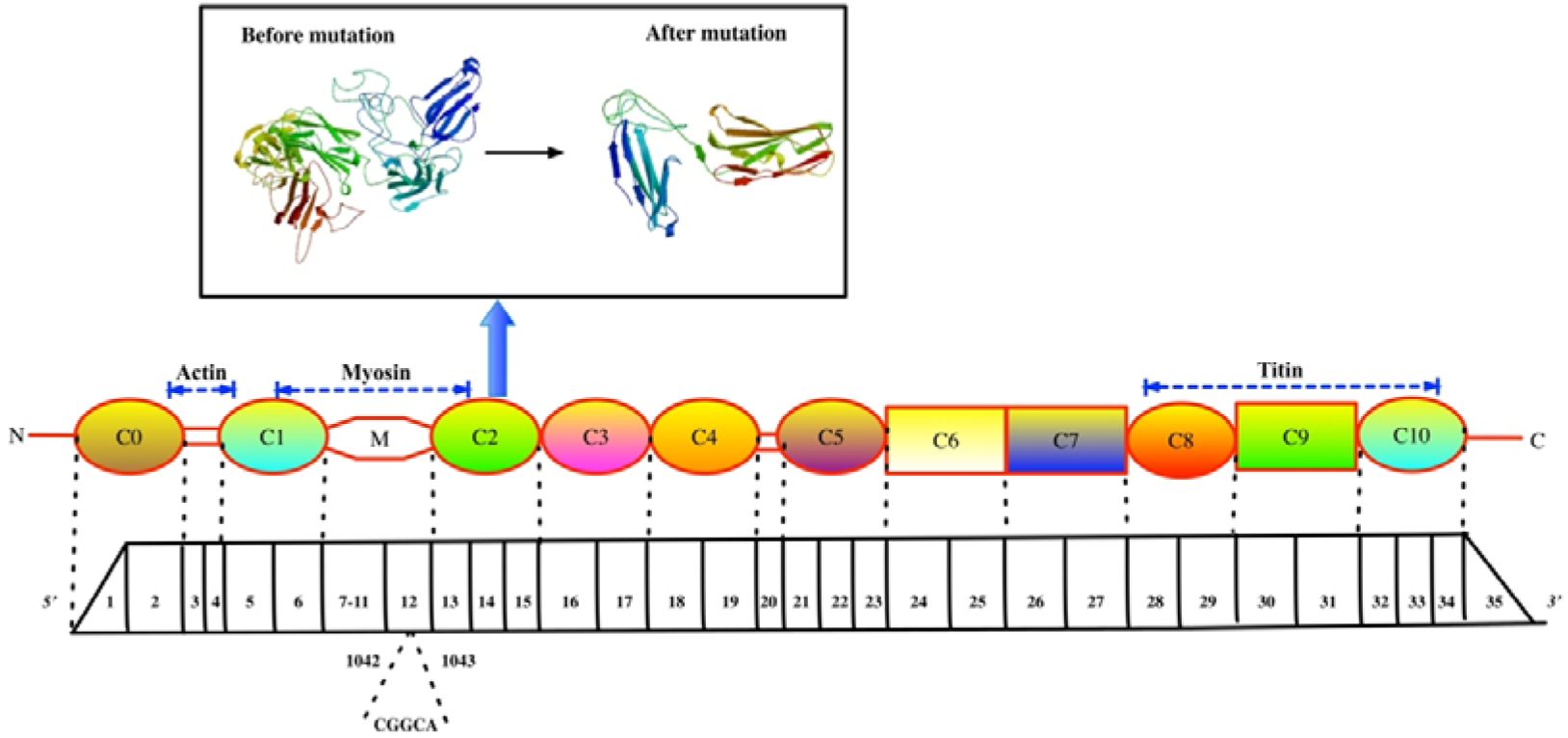
Structural representation of full-length cMyBP-C protein and MYBPC3 exons. Cartoon 3D model of cMyBP-C before and after mutation is shown in the rectangle. cMyBP-C is composed of 11 domains, being numbered C0 to C10 from the N-terminus to C-terminus. It includes 8 immunoglobulin (IgI) domains (circles) and 3 fibronectin type III (FnIII) domains (rectangles). Four specific phosphorylation sites located between C1 and C2, known as MyBP-C motif. The regions of interaction with other sarcomeric proteins (actin, myosin, and titin) is shown with dotted arrows. Relationship between the 35 exons of MYBPC3 cDNA and cMyBP-C domains is indicated by dashed lines.

## **4.** Discussion

### 4.1 Main findings

In this study, we described a novel mutation in the encoding region of MYBPC3 from a HCM family. All the four patients turned out to be compound heterozygote carriers of MYBPC3 gene. The mutation led to truncated MYBPC3 protein that lack the CLterminus myosin and titin binding domain. Although prior studies have reported HCM cases with MYBPC3 mutation[14], to the best of our knowledge, our study is the first to report the c.1042_1043insCGGCA mutation in exon 12.

### 4.2 Role of mutated MYBPC3 in hypertrophic cardiomyopathy

Variants in sarcomere and sarcomere-associated protein genes including MYBPC3, myosin heavy chain [MYH7], cardiac troponin T [TNNT2], cardiac troponin I [TNNI3], α-tropomyosin [TPM1], myosin essential and regulatory light chains [MLY2, MYL3], and actin [ACTC] are identified as the most common cause of HCM[15]. Mutations in MYBPC3 often lead to reduced expression of full length cardiac myosin binding protein-C (cMYBP-C), which is composed of a chain of eleven globular immunoglobulin and fibronectin domains (C0–C10) and an extensible M-domain between C1 and C2[16]. As in the present study, the frame shift mutation led to an early termination of cMyBP-C protein synthesis at C2 domain, disturbing its normal structural and regulatory functions.

### 4.3 Relationship between genotype and phenotype

Mutations in genes encoding contractile proteins result in contractile dysfunction of the cardiac fibers, leading to compensatory hypertrophy and decreased diastolic function[17]. Meanwhile, some gene mutation can interfere with calcium circulation and sensitivity of calcium ion in the myocardium, thus affecting its energy metabolism and leading to myocardial hypertrophy[17–19]. For different gene mutations, the functional mode or dominant effect of the gene products may be various, causing diverse phenotypes[15,20,21].

As in the present study, variant in MYBPC3 is the main contributor to the development of HCM. Studies have shown that mutations in MYBPC3 are relatively more benign when comparing with those in MYH7[15,22]. In addition, it was observed that patients with MYBPC3 variants often have an older age of onset, along with slower progression and thus better prognosis[23,24]. The proband in the current pedigree is consistent with a normal life expectancy. However, as in the case of individual II-1, sudden death may occur under extreme circumstances. Furthermore, when MYBPC3 is combined with other mutations, the phenotype can be much more serious.

### 4.4 Value of genetic testing

Genetic testing enables an early diagnosis among relatives of patients with hypertrophic cardiomyopathy[25]. It is of great significance for the differential diagnosis of diseases with similar clinical phenotypes and it may aid in risk stratification and helpful to establish individualized therapeutic strategies[26]. Physicians can provide accurate genetic counseling and prenatal testing with a genetic diagnosis[27]. As in the current study, HCM family members can benefit from the above-mentioned advantages of genetic testing. The development of sequencing technology contributes significantly to the improvement of screening, genetic counseling and risk stratification of HCM, holding promise for the prevention and early detection of the disease.

### 4.5 Conclusion

In conclusion, we performed next generation sequencing to identify a novel frameshift mutation mapped to MYBPC3 c.1042_1043insCGGCA, which was thought to be associated with late-onset myocardial hypertrophy and had not been reported before. Premature termination at C2 domain was predicted by computational tools and structural and functional disruption of cMyBP-C occurs, thereby leading to an abnormal, truncated protein or loss of protein from this allele due to nonsense-mediated mRNA decay. These features may provide important information for physicians in clinical management of hypertrophic cardiomyopathy. Further researches of the mutation c.1042_1043insCGGCA are needed.

## Data Availability

All relevant data are within the manuscript and its Supporting Information files.

## Acknowledgments

This work was supported by grants from Zhejiang Provincial Traditional Chinese Medicine Science and Technology Plan (2023ZF121) and Clinical Medical Research Project of Zhejiang Medical Association (2023ZYC-A05).

